# Impact of Adjustment for Differential Testing by Age and Sex on Apparent Epidemiology of SARS-CoV-2 Infection in Ontario, Canada

**DOI:** 10.1101/2023.08.01.23293449

**Authors:** Savana Bosco, Amy Peng, Ashleigh R. Tuite, Alison Simmons, David N. Fisman

## Abstract

**Background:** Surveillance of communicable diseases typically relies on case counts for estimates of risk, and counts can be strongly influenced by testing rates. In the Canadian province of Ontario, testing rates varied markedly by age, sex, geography and time over the course of the SARS-CoV-2 pandemic. We applied a standardization-based approach to test-adjustment to better understand pandemic dynamics from 2020 to 2022, and to better understand when test-adjustment is necessary for accurate estimation of risk.

**Methods:** SARS-CoV-2 case counts by age, sex, public health unit and week were obtained from Ontario’s Case and Contact Management system (CCM), which includes all SARS-CoV-2 cases from March 2020 to August 2022. Complete data on testing volumes was obtained from the Ontario Laboratory Information System (OLIS). Case counts were adjusted for under-testing using a previously published standardization-based approach that estimates case numbers that would have been expected if the entire population was tested at the same rate as most-tested age and sex groups. Logistic regression was used to identify threshold testing rates beyond which test-adjustment was unnecessary.

**Results:** Testing rates varied markedly by age, sex, public health unit and pandemic wave. After adjustment for under-testing, overall case counts increased threefold. Adjusted epidemic curves suggested, in contrast to reported case counts, that the first two pandemic waves were equivalent in size, and that there were three distinct pandemic waves in 2022, due to the emergence of Omicron variants. Under-reporting was greatest in children and young males, and varied significantly across public health units, with variation explained partly by testing rates and prevalence of multigenerational households. Test adjustment resulted in little change in the epidemic curve during pandemic waves when testing rates were highest; we found that test-adjustment did not increase case counts once weekly per capita testing rates exceeded 6.3%.

**Conclusions:** Standardization-based adjustment for differential testing by age and sex, and for dynamic changes in testing over time, results in a different picture of infection risk during the SARS-CoV-2 pandemic in Ontario; test-adjusted epidemic curves are concordant with observed patterns of mortality during the pandemic and have face validity. This methodology offers an alternative to sero-epidemiology for identification of true burden of infection when reinfection, sero-reversion, and non-specificity of serological assays make sero-epidemiology challenging.

## Introduction

The 2019 emergence of SARS-CoV-2 resulted in a global pandemic with severe impacts on mortality, life expectancy, population health status, and economies (1–9). While the pandemic *qua* pandemic appears to have subsided, this highly virulent airborne pathogen remains endemic worldwide. The true impact of the pandemic on population health continues to be debated, as does the likely impact of the virus’ ongoing endemic circulation. Understanding the pathogen’s impact depends on a clear vision of disease epidemiology, which in turn depends on accurate analysis of public health surveillance data.

Early in the pandemic we observed that the estimated incidence of SARS-CoV-2 infection in Ontario, Canada, was strongly predicted by frequency of PCR testing; in other words, groups such as older adults, who were tested more intensively, appeared to have higher SARS-CoV-2 incidence than groups tested less frequently (such as young males and children of both sexes) (10). We developed a simple, regression-based approach that permitted adjustment for testing frequency, such that it became possible to estimate the incidence of infection that *would have* been observed in a given age/sex group had it been tested at the same rates as the most highly tested population group. We found that adjusting for test frequency resulted in a very different view of the pandemic; one in which younger individuals (and in particular, males aged 20-29) represented a far larger share of infections than was observable in unadjusted data (10).

Our earlier analysis was restricted to the period from initial SARS-CoV-2 emergence in Ontario (which we dated to March 2020, when community transmission was first recognized) to December 2020 (10). However, this early period preceded the widespread emergence of novel viral variants of concern (VOC) and use of vaccination to control the pandemic. Indeed, Mitchell et al. have divided Canada’s SARS-CoV-2 into six distinct periods based on dominant circulating VOC and disease incidence, an approach which also partially captures the timing of SARS-CoV-2 vaccine roll-out in Canada (11).

Our objectives were: to extend our earlier analysis, to evaluate the differences between epidemic curves generated using unadjusted and test-adjusted case counts, both for the pandemic overall and for distinct, individual waves that occurred between March 2020 and September 2022; and to generate age- and sex-specific epidemic curves, to quantify the degree to which cases are likely to have been under-recognized due to under-testing in different age and sex groups. We also sought to identify testing thresholds above which test adjustment made little difference to perceived epidemic activity.

### Data Sources

We evaluated disease incidence using population-based SARS-CoV-2 infection data from the Ontario Case and Contact Management System (CCM), a data system used by Ontario’s 34 public health units for public health management of notifiable diseases (10). The case definition for SARS-CoV-2 during the period under evaluation required a positive nucleic acid amplification test assay (e.g., real time PCR) from an accredited laboratory (12). CCM included data on age (10-year intervals) and sex of case patients and date a positive SARS-CoV-2 PCR was reported; as this last data element was complete, we used it as a surrogate for case date (10). Laboratory testing volumes were obtained from the Ontario Laboratories Information System (OLIS), which includes testing and reporting dates for all PCR tests performed in the province, including tests performed in the public health laboratory system, hospital system and private laboratories. As such OLIS is believed to be a complete record of SARS-CoV-2 PCR testing in Ontario during the period under study. We counted the first test record when a person had multiple tests on a given day; however, subsequent testing of that person could be incorporated into test counts (10).

### Classification of Waves

We classified the SARS-CoV-2 pandemic in Ontario by slightly modifying the approach of Mitchell et al., who identified six distinct waves based on disease activity and dominant circulating viral variant of concern: waves 1 and 2 (wild-type dominant, with wave 1 ending on August 31, and wave 2 from September 1 2020 to February 28, 2021; wave 3 (mixed Alpha/Beta/Gamma variants, March 1 to June 30, 2021); wave 4 (Delta variant dominant, from July 1 to December 25, 2021); and Omicron dominant waves 5 and 6 (from December 26 2021 to March 19, 2022, and after March 20, 2022, respectively) (11). For each of these waves, we evaluated overall per capita testing by age and sex, and identified the most tested group during each wave.

### Adjustment for Under-testing

We used meta-regression-based methods to adjust case counts in other age and sex groups for under-testing, estimating the case rates that would have been expected if these groups were tested at the same rate the most tested group. This method is described in detail elsewhere (10), but briefly requires that a standardized infection ratio (SIR), and standardized testing ratio (STR), be estimated weekly, by public health unit, for each age- and sex-group, with incidence and testing rates in the most tested group used in the denominator of these ratios. For most waves, the most tested group was females aged 80 and over; however, for wave 4, the most tested group was male children under 10 years of age, and this group was used as the referent for that period.

Using wave-specific SIR and STR, it is possible to create age- and sex-specific meta-regression models using log-transformed of *i* age and sex groups, E(ln(SIR_i_)) = α + β(ln(STR_i_)). As ln(STR_i_) is zero when a given age and sex group is tested at the same rate as the most tested group, the model intercept α can be interpreted as the standardized infection ratio that would be expected in the presence of equal testing. This SIR can then be multiplied by observed infection incidence in the most tested group, to generate an estimate of test-adjusted incidence. Weekly test-adjusted case estimates by age and sex for each public health unit were generated in this way. Overall test-adjusted epidemic curves were created by summing adjusted case numbers.

### Analysis

We compared the crude epidemic curve for Ontario, and test-adjusted epidemic curves graphically, and we also evaluated the ratio of reported cases to test-adjusted case estimates by age group, sex, public health unit, and time period. We denoted this ratio a “reporting ratio” (denoted ReR to avoid confusion with “relative risk”). As overall ReR were < 1 (i.e., test-adjusted cases exceeded reported cases we calculated the standard error of ln(URR) as the square root of ((1/reported cases)+(1/(test-adjusted cases - reported cases))).

We quantified the relative magnitude of ReR by age, sex, period and public health unit, and evaluated statistical significance for differences between groups, using negative binomial regression models, using reported cases as the dependent variable, and adjusted case estimates as model offsets. Models included linear, quadratic and cubic time trend terms, as well as fast Fourier transforms (FFT) to capture disease seasonality. Age groups were treated as indicator variables, with the oldest age group (80+) used as the referent. Public health units were also included as indicator variables, with the public health unit with the median ReR (Kingston) used as the referent. Interaction between age and sex was evaluated using multiplicative interaction terms. Such a model produces “incidence rate ratios” which can be interpreted as a *relative* ReR (RReR) by age, sex, period, etc.

Heterogeneity in reporting ratios across public health units was evaluated using meta-analytic methods, with meta-regression used to quantify the extent to which such heterogeneity could be explained by variation in test rates, health resources, vaccine uptake, urbanicity, and socioeconomic and demographic characteristics of public health units. We used non-long-term care hospital beds per capita for each public health unit as a surrogate for local health resources; these were obtained from the Canadian Institute for Health Information (13). Vaccination coverage was defined as total cumulative SARS-CoV-2 vaccine doses per capita over the period under study; vaccination data were obtained from COVAXON as described elsewhere (14, 15). Urbanicity effect was evaluated by comparing effects within the “Greater Toronto/Hamilton Area” (GTHA), the province’s largest population concentration, to effects outside the GTHA. Socioeconomic and demographic characteristics of each public health unit were obtained from Statistics Canada’s 2021 Census data (16); these included mean population age; proportion of population over age 64; proportion of multigenerational households; income inequality (Gini coefficient based on after tax household income); percent visible minorities; percent of residents identifying as Indigenous; percent of residents who are new immigrants; percent of residents who own a home; unemployment rate; proportion of residents with low income; median after tax household income; proportion of residents with educational attainment less than high school graduation; and proportion of residents who are Canadian citizens. The associations between each of these public health unit-level characteristics and reporting ratios were evaluated with univariable meta-regression models; factors with P ≤ 0.20 were evaluated in a multivariable meta-regression model built using backwards elimination.

We noted that although adjusted case counts were, overall, higher than crude case counts, test adjustment resulted in little change in the overall epidemic curve during periods when testing rates were high, and in some age- and sex-groups, and time periods adjusted case counts were, in fact, lower than crude reported case counts. We evaluated the relationship between this phenomenon (test adjustment resulting in *decreased* cases counts) and group-specific testing rates using logistic regression models. All analyses were performed in Stata version 15. The study was approved by the Research Ethics Board of the University of Toronto.

## Results

Between March 1, 2020 and September 4, 2022, 22.82 million PCR tests were performed for SARS-CoV-2 in the province of Ontario with 1,377,146 (6.03%) of these tests positive. Testing rates varied over time, with two distinct peaks: from December 2020 to March 2021, when per capita test rates were above 2% per week; and in December 2021 and January 2022, when per capita testing rates approached 3% per week (**Figure 1.A.**). Females aged 80 and over were the most-tested group over the study time period and were tested at a rate of 2.7% per week; males aged 10-19 were tested at the lowest rates (0.7% per week) (**Figure 1.B.**). When we evaluated testing by age and sex for each pandemic wave, we found that for 5 of 6 identified waves, females aged 80 and over were the most tested group; however, for wave 4 (Delta-variant dominant) the most tested group was males aged < 10 years (**Figure 1.C.**).

**Figure 1.**
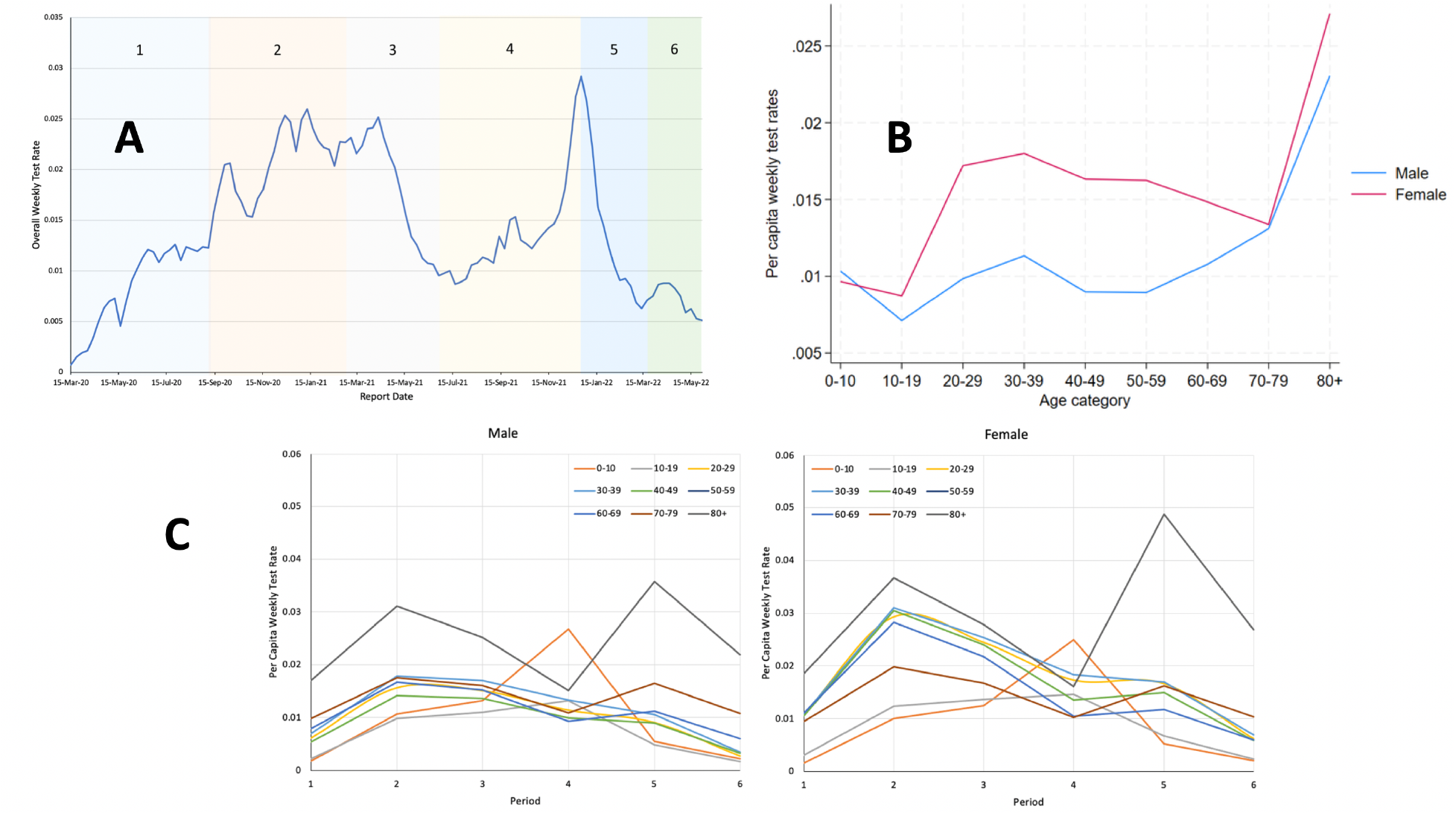
Weekly Rates of Testing for SARS-CoV-2 in Ontario, Canada, March 2020 to August 2022. (A) Overall weekly per capita test rates by date of test report. Shading and numbers denote distinct pandemic waves as defined by Mitchell et al. (11). (B) Average per capita weekly test rates by age group for males and females over the entire study period. (C) Average per capita weekly test rates for males (left panel) and females (right panel) by pandemic wave. Colors of curves correspond to individual age groupings; for all waves except wave 4 the most tested group is females aged 80 and over; in wave 4 the most tested group was 0- to 10-year-old males.

After adjustment for under-testing based on most-tested group in each wave, we were able to construct a test-adjusted epidemic curve and compare it graphically to the reported epidemic curve. Test adjustment resulted in a different appearance to the epidemic curve at the beginning and end of the pandemic, when testing per capita was at its lowest (**Figure 2.A.**). In particular, the first wave of the pandemic in spring 2020, and the second wave that autumn, appeared equivalent in magnitude after test adjustment. Test adjustment also markedly increased the apparent magnitude of the first wave caused by Omicron variant emergence and identified two subsequent large waves of infection caused by Omicron in spring and summer of 2022. By contrast, test-adjusted and unadjusted curves, for the mid-pandemic period when testing was most intense, were almost identical. Graphical comparison of the test adjusted epidemic curve to a plot of SARS-CoV-2 attributed deaths by week appeared to provide validation for the test-adjusted epidemic curve, with almost identical magnitudes in spring and autumn 2020 (waves 1 and 2 respectively), and three distinct waves of mortality after Omicron variant emergence (**Figure 2B**).

**Figure 2.**
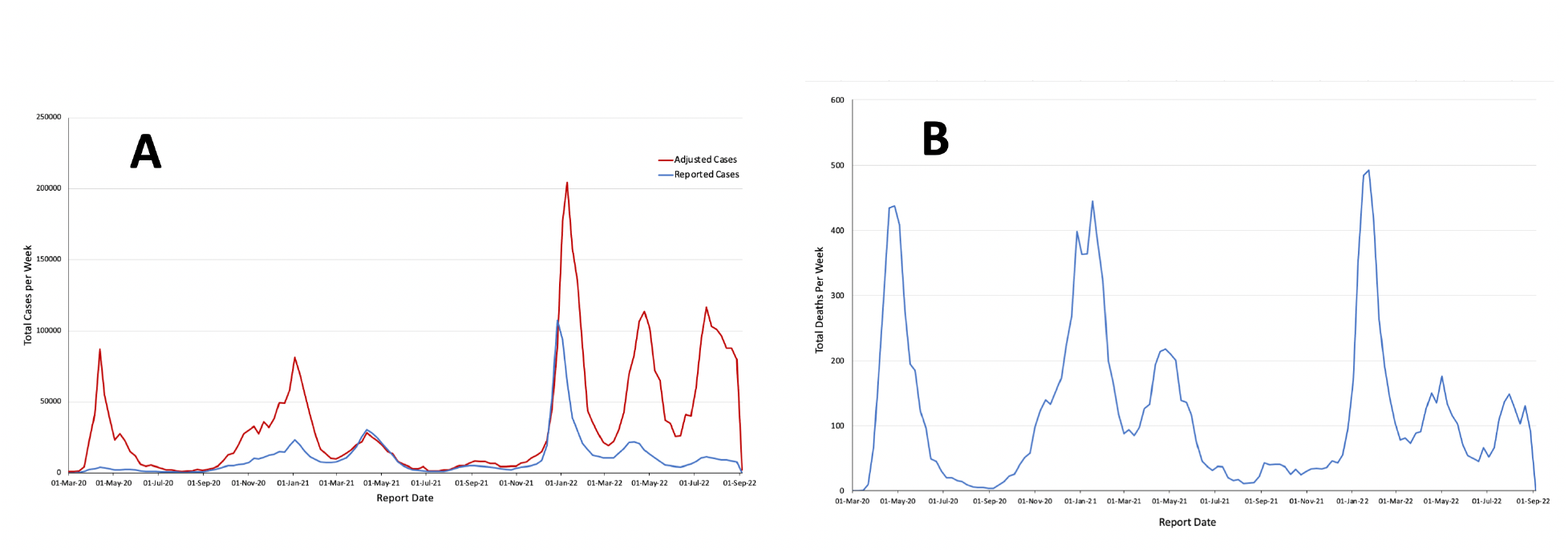
Reported and Test-Adjusted Epidemic Curves, and Correspondence with SARS-CoV-2 Mortality in Ontario, Canada. (A) Reported (blue) and test-adjusted (red) case counts over the course of the SARS-CoV-2 pandemic. Early and late pandemic waves are far larger with test adjustment, reflecting low overall rates of testing. (B) SARS-CoV-2 deaths in Ontario over time. Similar numbers of deaths in the first and second waves, and three distinct waves of deaths with emergence of Omicron variants at the end of the pandemic, correspond with patterns seen in the test-adjusted epidemic curve, but not with patterns seen in the reported case epidemic curve.

Directly calculated ReR by age, sex, and time-period are presented graphically in **Appendix Figure 1**; ReR were highest in females aged 20-59 but below 1 in all age and sex groups. ReR approached 1 during the third and fourth pandemic waves. Negative binomial models identified significant (P < 0.001) interaction between age group and sex in relative under-reporting, with relative ReR higher (i.e., closer to 1) in females than in males. Patterns of relative reporting, adjusted for age, sex, period and geography, were similar in negative binomial models to patterns identified via calculation of crude ReR (**Table 1**).

**Table 1.** Adjusted Relative Reporting Ratios for SARS-CoV-2 by Age, Sex and Time Period. Estimates are derived from a negative binomial regression model, with reported case counts used as dependent variable and test-adjusted counts used as model offsets. Models are also adjusted for public health unit and time trends. Male and female estimates are presented separately due to significant interaction between age and sex.

| **Covariate** | **Adjusted**<br>**Relative**<br>**Reporting**<br>**Ratio** | **95%**<br>**Confidence**<br>**Intervals** | **P-value** |
| --- | --- | --- | --- |
| **Age, Male** |  |  |  |
| 0 to 9 | 0.43 | 0.41-0.46 | <0.001 |
| 10 to 19 | 0.49 | 0.47-0.52 | <0.001 |
| 20 to 29 | 0.84 | 0.80-0.88 | <0.001 |
| 30 to 39 | 0.88 | 0.84-0.92 | <0.001 |
| 40 to 49 | 0.79 | 0.75-0.83 | <0.001 |
| 50 to 59 | 0.79 | 0.75-0.83 | <0.001 |
| 60 to 69 | 0.82 | 0.78-0.86 | <0.001 |
| 70 to 79 | 0.78 | 0.74-0.82 | <0.001 |
| 80 and over | 1.06 | 1.01-1.12 | <0.001 |
| **Age, Female** |  |  |  |
| 0 to 9 | 0.48 | 0.45-50 | <0.001 |
| 10 to 19 | 0.61 | 0.58-0.64 | <0.001 |
| 20 to 29 | 1.35 | 1.29-1.41 | <0.001 |
| 30 to 39 | 1.33 | 1.27-1.40 | <0.001 |
| 40 to 49 | 1.20 | 1.15-1.26 | <0.001 |
| 50 to 59 | 1.07 | 1.02-1.12 | 0.006 |
| 60 to 69 | 0.91 | 0.87-0.96 | <0.001 |
| 70 to 79 | 0.68 | 0.65-0.71 | <0.001 |
| 80 and over (referent) | 1.00 | --- | --- |
| Time Period (Pandemic Wave) |  |  |  |
| 1 (referent) | 1.00 | --- | --- |
| 2 | 0.86 | 0.81-0.92 | <0.001 |
| 3 | 1.43 | 1.32-1.56 | <0.001 |
| 4 | 1.30 | 1.15-1.48 | <0.001 |
| 5 | 1.05 | 0.92-1.20 | 0.488 |
| 6 | 0.58 | 0.50-0.67 | <0.001 |

Overall, ReR across public health units ranged from 0.11 (in Timiskaming) to 0.69 (in Peel); the overall pooled ReR was 0.31, with significant heterogeneity (P < 0.001) (**Figure 3** and **Appendix Figure 2**). In univariable meta-regression models, testing rate, fraction of multigenerational households, percent of population identifying as Indigenous, and non-long term care hospital beds per capita were each associated with URR with P ≤ 0.20. Only testing rate and multi-generational households remained in our final multivariable meta-regression model due to our use of backwards elimination. We found that each percentage increase in the proportion of the population tested resulted in a 2.55-fold increase in the ReR, while each percentage increase in multigenerational households resulted in a 1.08-fold increase in ReR (**Table 2** and **Appendix Figure 3**).

**Figure 3.**
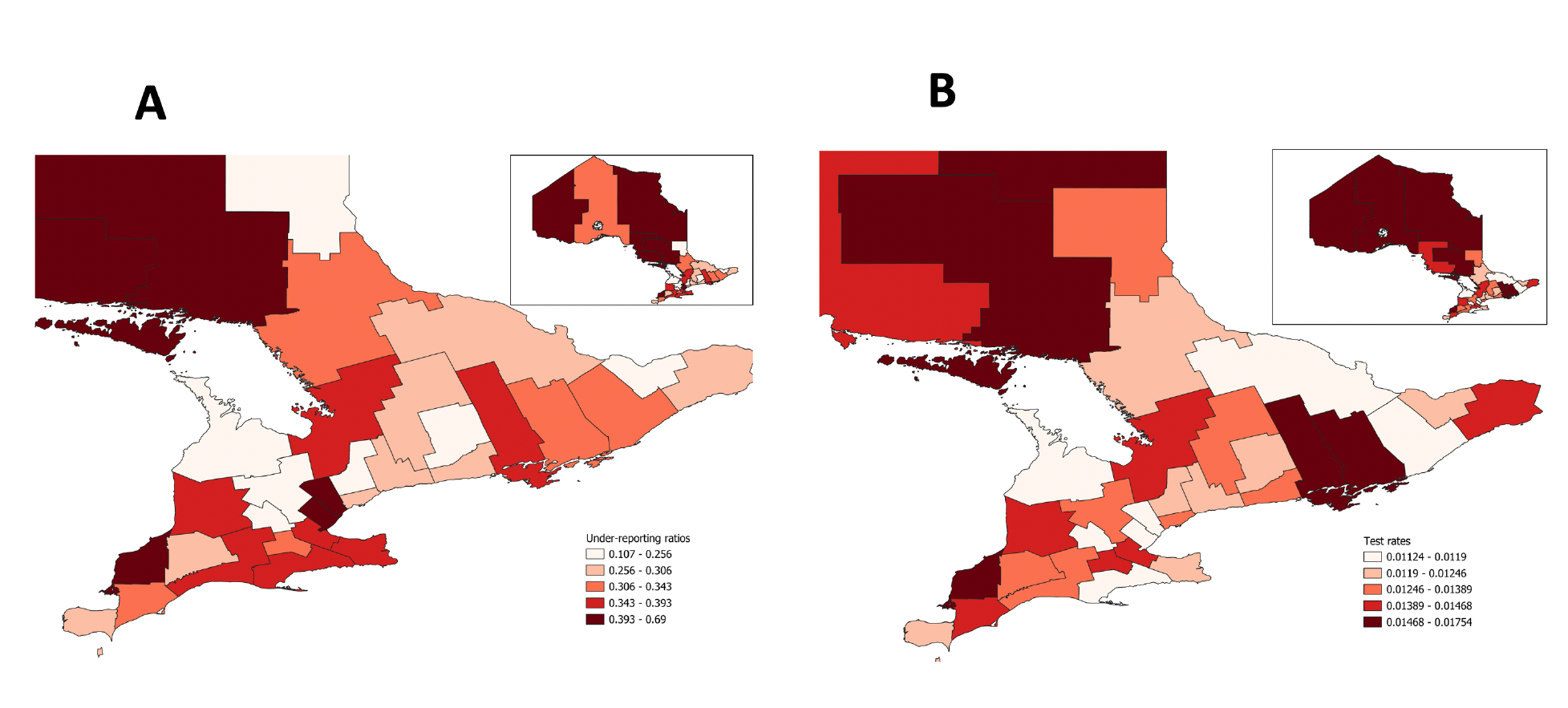
Reporting Ratios and Average Weekly Testing Rates in Ontario Public Health Units. (A) Reporting ratios by health unit in Southern Ontario (entire province is presented in inset). Higher (darker) regions have higher reporting ratios, meaning that unadjusted case counts are relatively closer to test-adjusted case counts than in other health units. Lighter regions have lower reporting ratios, indicating a larger relative gap between reported case counts and test-adjusted case counts. (B) Average weekly SARS-CoV-2 testing rates by health unit. Darker regions had higher rates of testing.

**Table 2.**
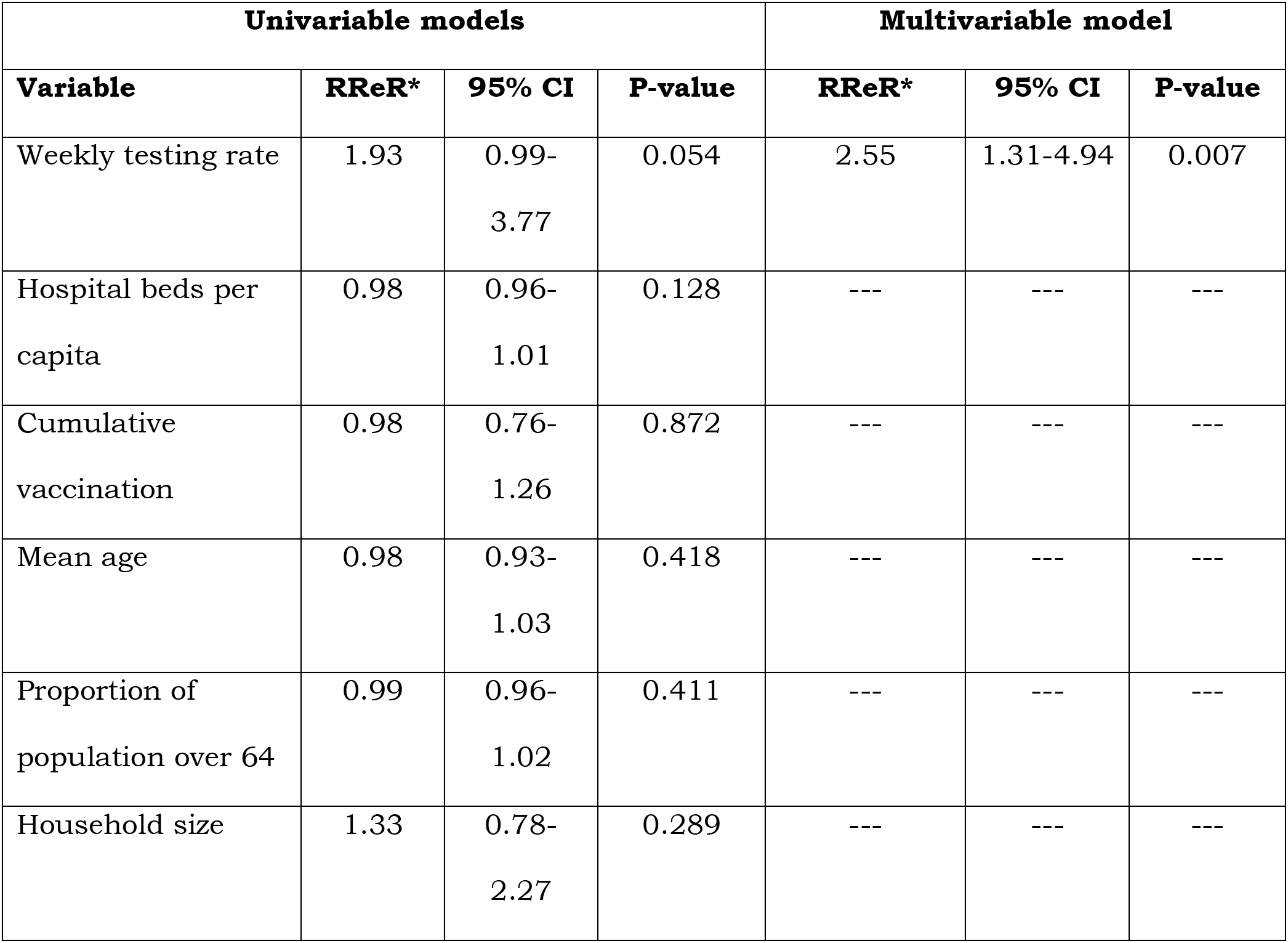
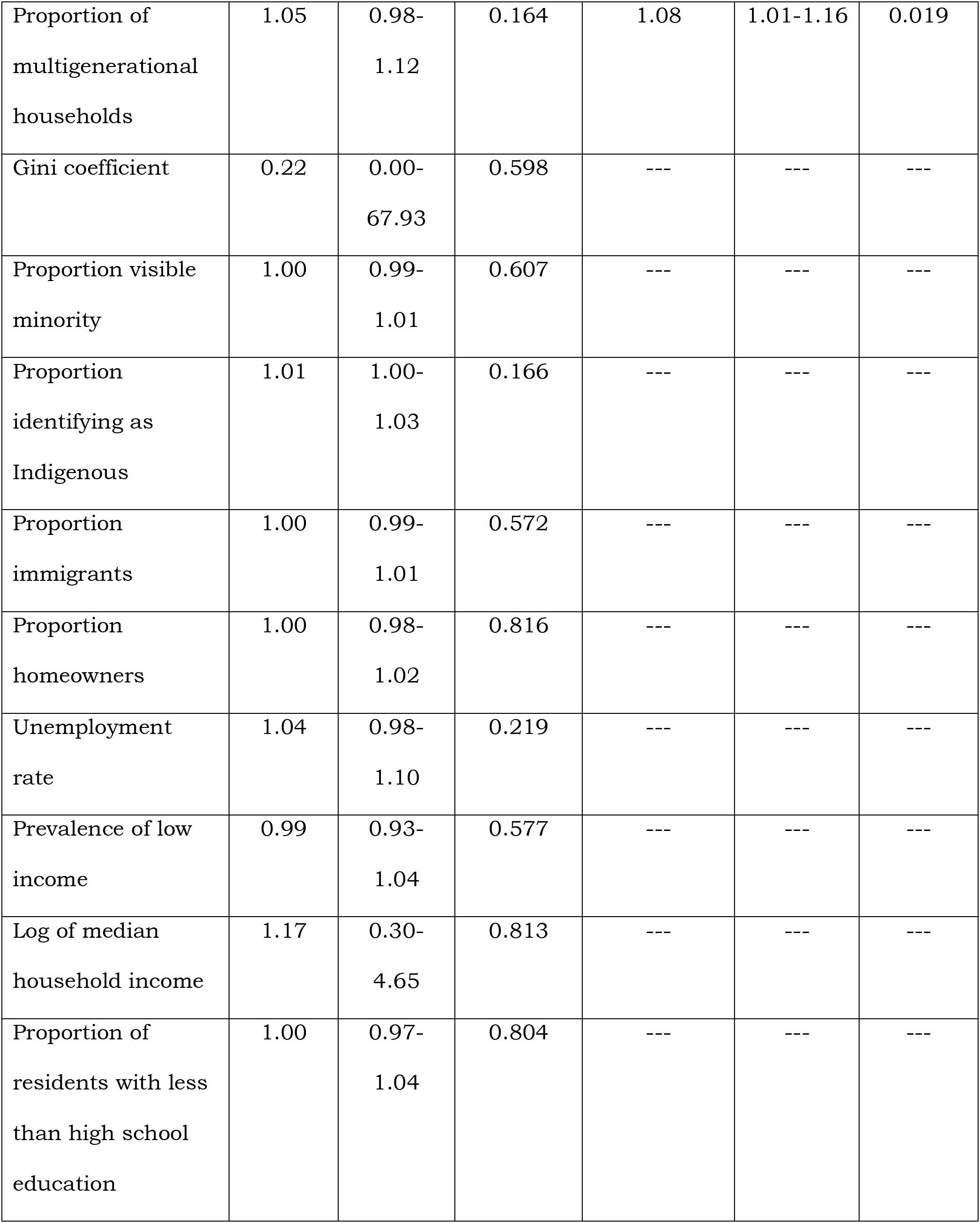
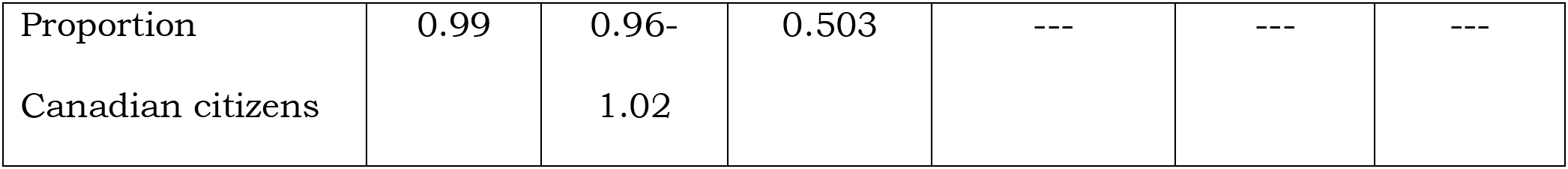
Univariable and Multivariable Meta-regression Models of Variation in Relative Reporting Ratio by Public Health Unit. Predictors with P-values less than or equal to 0.20 in univariable meta-regression models were included as candidate predictors in a final multivariable model, which was reduced using backwards elimination; only weekly testing rate and proportion of multigenerational households were retained in the final model.

We evaluated the relationship between adjusted case counts equal to, or lower than, crude reported case counts and testing rates using logistic regression. For every one percent increase in per capita weekly testing, the odds of adjusted case counts being less than or equal to crude case counts increased by 39% (95% CI 37% to 41%, P < 0.001). The weekly per capita test rate at which adjusted case counts were equally likely to be higher or lower than reported case counts was 6.26% (95% CI 6.08% to 6.47%) (**Figure 4**).

**Figure 4.**
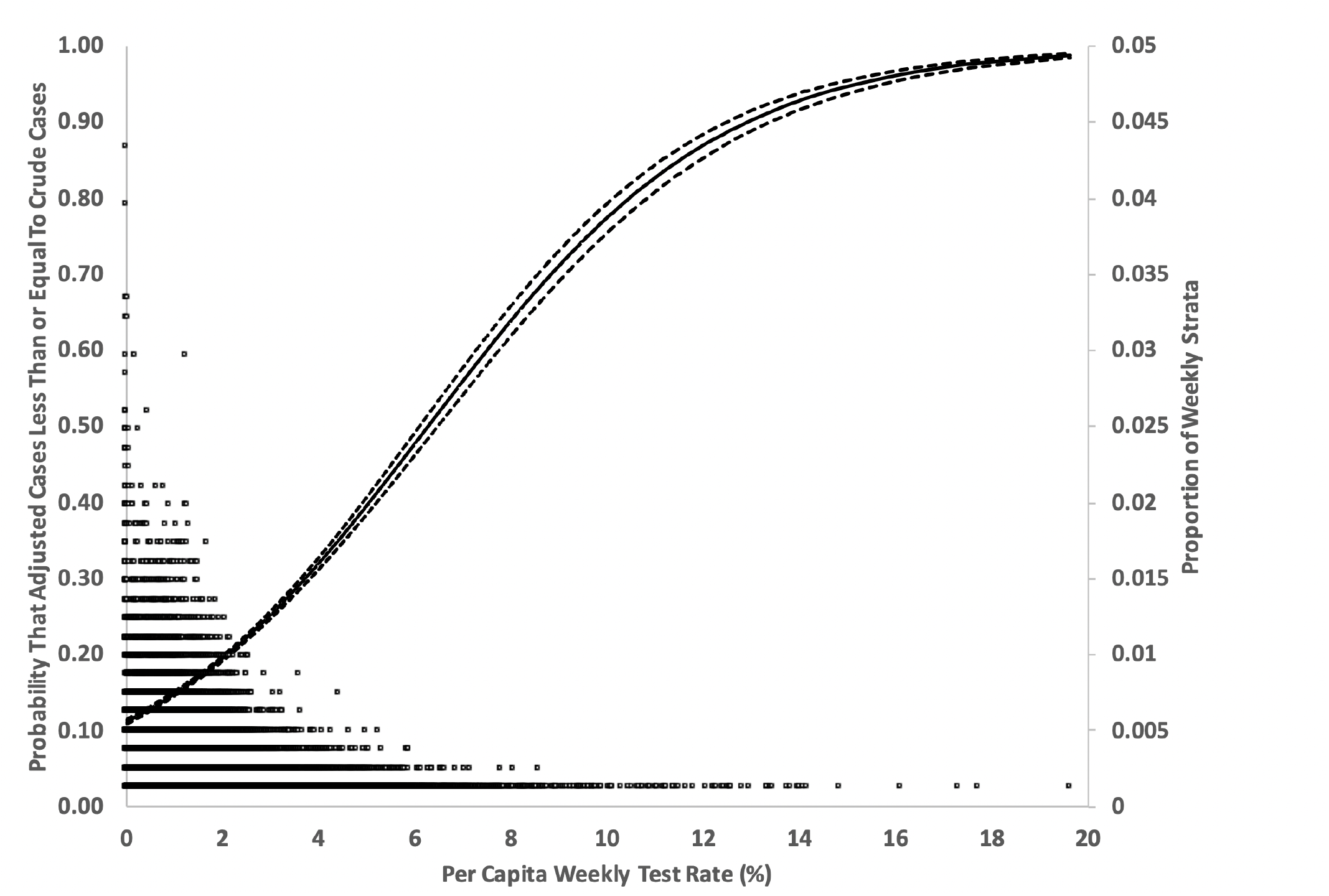
Testing Frequency and Likelihood of Test-Adjusted Case Counts Less Than or Equal To Reported Case Counts. Solid (point estimate) and dashed (95% confidence intervals) curves show the relationship between per capita weekly test rate (X-axis) and the probability that test-adjusted cases in a given stratum are less than or equal to reported case counts (left-hand Y-axis). If test adjustment does not increase case counts, test adjustment is not helpful in understanding disease epidemiology. It can be seen that when per capita testing rate exceeds 6.3% per adjusted case counts are more likely than not to be equivalent to, or less than, reported case counts. Circles show proportion of per capita weekly test rates (right sided Y-axis) by age, sex and public health unit strata, for each testing rate (X-axis). It can be seen that most testing during the study period was at rates below the threshold rate of 6.3% per week.

## Discussion

While case counts are the mainstay of public health surveillance systems, it is often forgotten that case counts are heavily influenced by testing volume. The tendency to focus testing on individuals and groups at highest risk for severe outcomes during the SARS-CoV-2 pandemic led us to develop a standardization-based approach to test adjustment, which captures non-linear relationships between testing rates and case rates, by indexing them to incidence in highly tested age and sex groups (10). In applying this method across the duration of the SARS-CoV-2 pandemic in Ontario, we find that test-adjustment results in a changed picture of risk during the pandemic, particularly during early and late pandemic periods when test volumes were low.

Notably, we found that the apparent differences in the magnitude of the first two pandemic waves likely reflected changing testing volumes rather than changing disease epidemiology, consistent with the similarity in mortality associated with each of these waves. Test adjustment also made it clear that Ontario experienced three distinct waves of illness due to Omicron variants (BA1, BA4, and BA5) from winter to summer of 2022. The latter two waves were obscured by falling testing rates from January 2022 onwards, but again, the test-adjusted epidemic curve is consistent with three distinct waves of SARS-CoV-2 death present in Ontario’s data.

We found that testing rates resulted in substantial heterogeneity in the extent to which cases were under-reported by age-group and sex. Overall, cases in females were more likely to be identified than cases in males, not only because of intensive testing in the oldest females who represented the majority population in the province’s long-term care facilities. We also found that there was less under-reporting in younger adult females (ages 20 to 59), which may reflect different health (17), but may also reflect the demographics of Ontario’s female-majority healthcare workforce. By contrast, reporting ratios were notably low in children and younger male adults over the study period, despite higher rates of testing in schools in pandemic wave 4, suggesting that infection was under-recognized in these groups.

We also found considerable heterogeneity in under-reporting by health units; this heterogeneity was partially explained by variation in testing rates across health units, but increased prevalence of multi-generational households in a health unit was also associated with a significant narrowing of the gap between reported and test-adjusted case counts. This finding has considerable face validity: if public health practice throughout the pandemic was to advise testing of household contacts of SARS-CoV-2 cases, occurrence of cases in multi-generational households would, by definition, have resulted in enhanced testing across age groups.

While we found that adjustment for under-testing created a different view of the pandemic than that apparent with reported cases, our analysis was also able to identify thresholds for population testing frequency beyond which adjustment for differential testing was not needed. During the middle of the pandemic, test rates were high, and we found little difference between epidemic curves generated using reported cases, and those generated using test-adjusted cases, in pandemic waves 3 and 4. This suggests an approach for more accurate evaluation of infection risk during epidemics or pandemics of novel infectious diseases, with test-adjustment applied during periods of under-testing, while crude case counts can be considered representative of disease epidemiology during periods of high testing.

Our approach represents an advance over traditional approaches to adjust for testing volumes in public health surveillance systems. This is typically achieved by incorporating test volumes as denominators, such that disease risk is presented based on percentage positivity of tests; the Canadian FluWatch system, which is a laboratory-based surveillance system for influenza, uses such an approach (18). While percentage positivity normalizes case counts for testing volumes, such an approach has limitations as an index of risk: the relationship between testing volume and test positivity is not fixed, but appears to be bidirectional (i.e., increasing risk or awareness of risk results in increased testing, while increased testing identifies increasing numbers of cases, albeit cases with varying risk profiles).

It might be argued that true understanding of the burden of infection (rather than disease) caused by SARS-CoV-2 should depend on sero-epidemiological methods rather than adjusted PCR testing. However, in the context of SARS-CoV-2 in Ontario, serological data have a number of important limitations, including (i) the lack of sensitivity and specificity of lone serological assays (19); (ii) dependence on potentially non-representative populations such as blood donors (19); (iii) loss of antibody positivity via seroreversion (19, 20); (iv) challenges in distinguishing seropositivity due to infection from seropositivity due to vaccination (19, 21); and (v) challenges associated with ubiquitous infection and repeated infection.

As with any observational research our work has limitations. In particular, we are not able to directly validate our test-adjusted case counts, given the limitations of sero-epidemiological data as noted above. However, the concordance of our test-adjusted waves with death waves during periods of under-reporting is suggestive of the validity of our approach. We are also not able to evaluate the generalizability of our approach outside Ontario or Canada, and are limited to the period prior to September 2022, as relevant data became unavailable after this time (22).

In summary, we applied a standardization-based approach to population-based case data from the SARS-CoV-2 pandemic in Ontario, Canada, and found that test-adjustment resulted in a different view of the pandemic during periods when testing rates were low. In particular, test adjustment was helpful in understanding the distribution of deaths over time by demonstrating that low case counts early and late in the pandemic represented under-testing rather than low incidence of infection. Further refinement of this approach by incorporation of testing data into public health surveillance may be possible through application to other disease processes (23) and in other jurisdictions.

## Appendix

**Appendix Figure 1.**
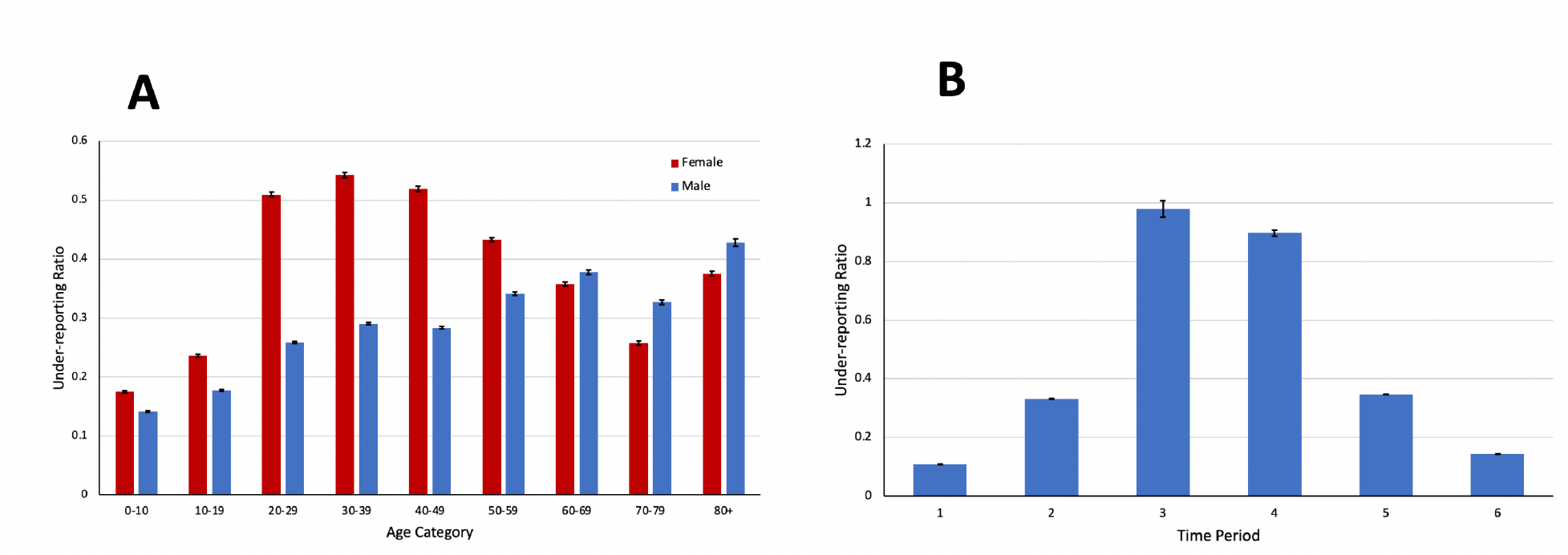
Reporting Ratios by Age, Sex and Time Period. Panel A (left) shows reporting ratios (Y-axis), defined as the ratio of reported cases to test-adjusted cases, over the study period by age-grouping (X-axis) and sex. Red bars represent females; blue bars represent males. Whiskers represent confidence bounds. Higher reporting ratios indicate a smaller relative gap between reported cases and test-adjusted cases. Panel B (right) shows reporting ratios for the population as a whole in each of six distinct pandemic waves. During waves 3 and 4, in the presence of high rates of testing, reporting ratios were close to 1.

**Appendix Figure 2.**
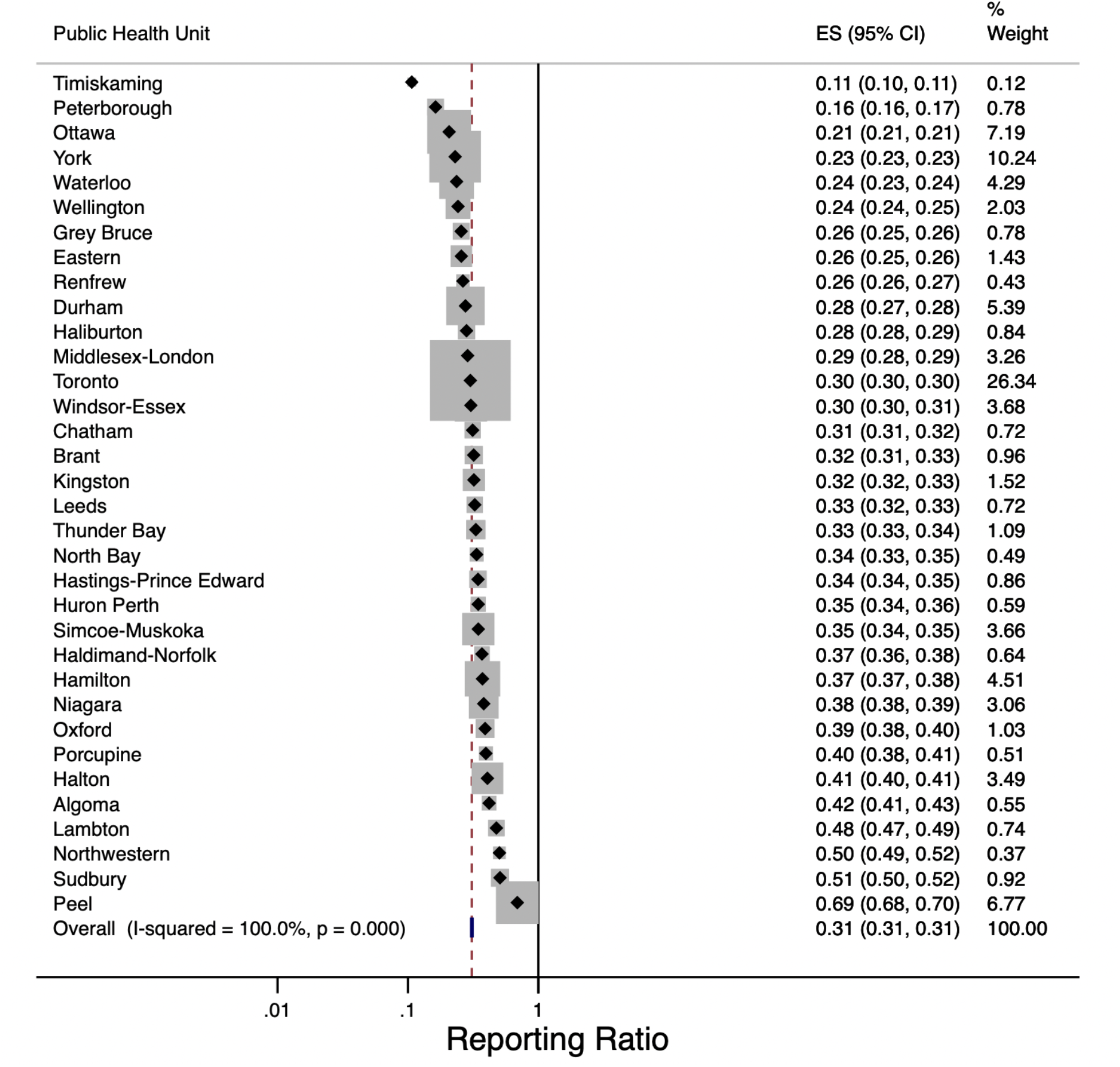
Forest Plot of Reporting Ratios by Ontario Health Unit. Health unit names are listed on the left size of the figure; reporting ratios (“effect size”, or “ES”) are listed on the right with 95% confidence intervals. Box sizes are inversely proportional to variance of estimates. Ratios range from 0.11 in Timiskaming to 0.69 in Peel, with a mean reporting ratio of 0.31 across all health units, and substantial between health unit heterogeneity.

**Appendix Figure 3.**
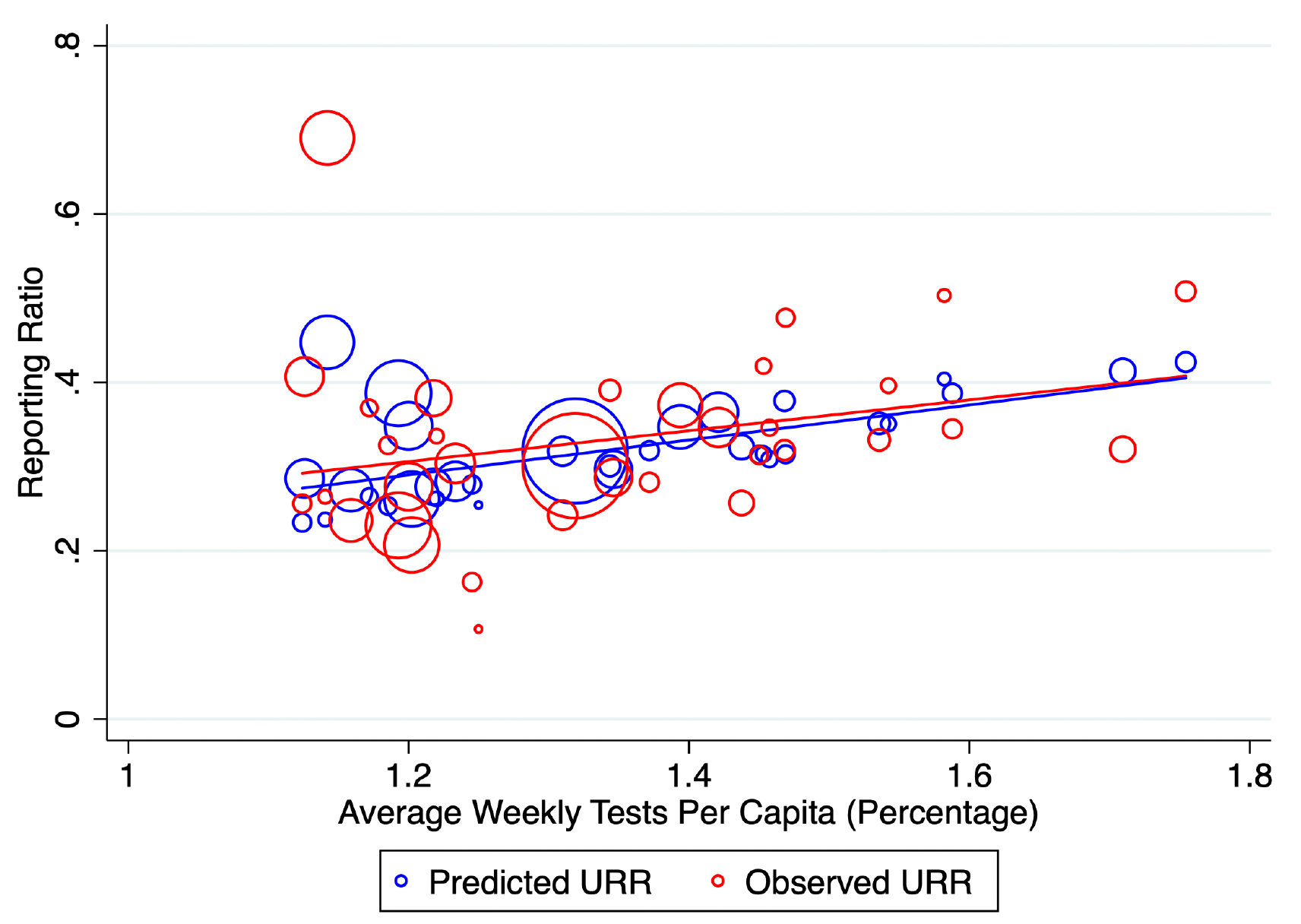
Observed and Predicted Reporting Ratios and Per Capita Testing. Circles represent observed (red) and predicted (blue) reporting ratios for SARS-CoV-2 by Ontario public health unit. Predicted reporting ratios are derived from a meta-regression model, which also adjusts for prevalence of multi-generational households by public health unit. Size of circles is inversely proportional to the variance of the estimate. Straight lines are lines of best fit for observed (red) and predicted (blue) estimates.

## Data Availability

The full individual-level dataset from which the analytic dataset is derived is the property of the Ontario Ministry of Health. The aggregate dataset used for this analysis can be obtained by contacting Dr. Fisman.

